# Visual and refractive status of children with Down’s syndrome and nystagmus

**DOI:** 10.1101/2021.07.17.21260703

**Authors:** Asma A A Zahidi, Lee McIlreavy, Jonathan T Erichsen, J Margaret Woodhouse

**Affiliations:** Optometry Programme, School of Health Professions, University of Plymouth, Plymouth, United Kingdom; School of Optometry and Vision Sciences, Cardiff University, Cardiff, United Kingdom; School of Optometry and Vision Sciences, Cardiff University, Maindy Road, Cathays, Cardiff CF24 4HQ

## Abstract

**Background/Aims:** Children with Down’s syndrome (DS) are known to have poorer visual acuity that neurotypical children. One report has shown that children with DS and nystagmus also have poor acuity when compared to typical children with nystagmus. What has not been established, is the extent of any acuity deficit due to nystagmus and whether nystagmus impacts on refractive error is within a population with DS.

**Methods:** Clinical records from The Cardiff University Down’s Syndrome Vision Research Unit were examined retrospectively. Binocular visual acuity and refraction data were available for 50 children who had DS and nystagmus (DSN) and 176 children who had DS but no nystagmus. Data were compared between the two groups, and with published data for neurotypical children with nystagmus.

**Results:** The study confirms the deficit in acuity in DS, compared to neurotypical children, of approximately 0.2 LogMAR and shows a further deficit attributable to nystagmus of a further 0.2 logMAR beyond the first year of life. Children with DS and no nystagmus appear to have acuity that mirrors that of typical children with nystagmus, while children with both DS and nystagmus have a significant additional impairment. Children with DS have a wide range of refractive errors, but nystagmus increases the likelihood of myopia. Prevalence and axis direction of astigmatism, on the other hand appears unaffected by nystagmus.

**Conclusion:** Nystagmus confers an additional visual impairment on children with Down’s syndrome and must be recognised as such by families and educators. Children with both DS and nystagmus clearly need targeted support.

## INTRODUCTION

Infantile nystagmus (IN) is one of the most frequently seen ocular disorders in children with Down’s syndrome (DS) and is estimated to occur in 15-30% of the population.(1)(2) Among neurotypical children, IN is associated with poorer visual acuity (e.g. Fu et al., 2011)(3) and a wide spectrum of refractive errors associated with failure in emmetropisation.(4)

There is little published data on visual acuity (VA) in children with DS and nystagmus (DSN). A study by Felius et al. (5) investigated the VA deficit of 16 children with DSN between the age of 10 months and 14 years using Teller cards. The VA reported was poorer than neurotypical children with nystagmus, but no comparison was made with children with DS and no nystagmus. It has been widely reported that children with DS have poorer visual acuity than the neurotypical norm. (6)(7)(8)(9)(10)(11) However, note that, in these studies, children with nystagmus or any other visually impairing condition are often excluded.(9)(10)(11)

The prevalence of refractive errors has been reported to be much higher in both children and adults with DS compared to the typical population.(12)(13) Refractive errors in infants with DS are similar to those of typical infants, but emmetropisation does not happen(14), so that the prevalence and degree of refractive errors increase and remain high.(15) To date, there are no published data concerning the refractive status of children with DSN exclusively. Therefore, information on the visual and refractive status of these children is very limited.

The aim of this study was to determine whether there are any differences in the distribution and development of VA and refractive error among children with DSN compared to those of children with DS, by analysing, retrospectively, the clinical records of children participating in the Cardiff University Down’s Syndrome Vision Research Unit (CDSVRU) studies.

## METHODS

Two hundred and fifty-eight clinical records of children in the CDSVRU between 1992 and 2017 were examined retrospectively. The recruitment procedures were explained in detail in Zahidi, Vinuela-Navarro and Woodhouse(11). Inclusion criterion was a diagnosis of Trisomy 21, and there were no exclusion criteria. Qualified optometrists conducted optometric assessments at the children’s home, school or in the clinic at the School of Optometry & Vision Sciences, Cardiff University (UK).

The method of visual acuity measurement varied, depending on the child’s age and cognitive ability, but was confined to preferential looking at 38cm with Teller Acuity cards (Precision Vision)(16) or at 50 cm with the Cardiff Acuity Test(17), Kay Picture LogMAR test (singles or crowded)(18), or Keeler LogMAR Crowded test(19), both at 3m. Depending on the child’s cooperation, VA was measured binocularly first, and then monocularly. Only binocular data were included in this analysis. Children who had been prescribed spectacles wore their corrections during VA measurement. This longitudinal study obtained continual and on-going approval from NHS Ethics in Wales (National Institute for Social Care and Health Research Ethics Service 08/MRE09/46, amendment 5, 7th July 2016). Study information was given to parents, and written consent was obtained from the parents of all participants involved. This study was conducted in accordance with the Declaration of Helsinki.

Refraction was performed using the Mohindra technique in a completely dark room or light-proof tent using a dim retinoscope light following the procedure outlined by Elliot(20). This technique has been shown to obtain results not significantly different from cycloplegic refraction in children with DS.(14) Refractive error was recorded in sphere, minus cylinder (cyl) form and axis. Significant refractive error was defined as spherical equivalent refractive error (SER) of <-0.50DS (myopia) or ≥+2.50DS (hypermetropia).(21) Significant astigmatism was defined as <-0.50DC. Data for the right eye (RE) were used for analysis for all participants except for those with anisometropia (a difference in SER ≥1.00D between right and left eyes) when the data of the least ametropic eye was used.(22) Axis of astigmatism was recorded as with-the-rule (minus cyl axis 180° ± 15°), against-the-rule (minus cyl axis 90° ± 15°) or oblique (minus cyl axis greater than ± 15º from the horizontal and vertical meridian).(21)

Children who presented with either manifest or latent nystagmus during two or more visits were identified and grouped into the Down’s syndrome with nystagmus (DSN) group, with the remainder in the non-nystagmus group (DS). Records from thirty-two children were excluded for the following reasons: 1) there were no visits in which binocular acuity data were obtained (n=8), 2) the age when entering the study was over 12 years old (n=13), 3) ocular condition such as cataract (n=2), and 4) not fully corrected during visual acuity measurement (n=9).

To prevent any bias, the database was inspected without names (codes were used) or acuity and refractive error data. The children were allocated to 7 age groups to enable meaningful comparison of the findings with that of typically developing children with nystagmus (3)(4)(23)(24): 1-11.9 months, 12-23.9 months, 2-3.9 years, 4-5.9 years, 6-7.9 years, 8-9.9 years, 10-11.9 years. Participants were limited to inclusion in one age group only.

Statistical analysis was performed using the IBM SPSS version 23 statistical package. Descriptive analysis was performed on both VA and refractive error data to determine the mean, standard deviation (SD), median, 95% confidence intervals, and frequency of each age group for both the DSN and DS group. The distribution of binocular VA, SER and astigmatism data for each group of children at each age group was tested for normality using the Shapiro-Wilk test. Data that were normally distributed (p>0.05) were analysed using parametric statistical tests, otherwise non-parametric statistical tests were used.

## RESULTS

After exclusions, 226 children were included in the cross-sectional study, which consisted of children with DSN (n=50) and DS (n=176). Of these, 91(40%) were female and 135(60%) were male. [Note that the acuity of 159 children in the DS group has already been reported(11). The database was updated for the current analysis and age groups modified]. The distribution of children in each age group is presented in Table 1.

**Table 1.**
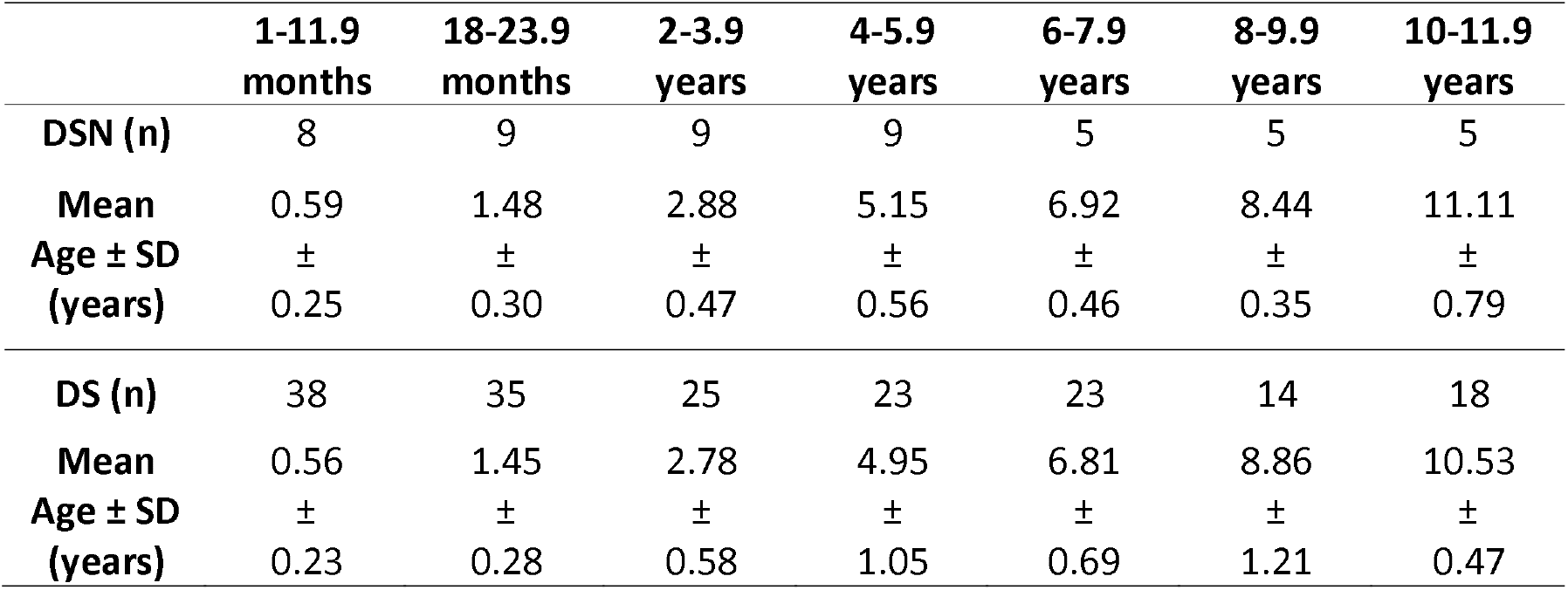
Distribution and mean age ± SD of children with DS and nystagmus (DSN) and without nystagmus (DS) in each age group

### Visual acuity

The mean binocular acuity (BVA) of children with DS and DSN is shown for age groups in Figure 1. BVA ranged from 0.2 and 1.4 LogMAR for the children in the DSN group and 0.0 and 1.4 LogMAR for the children in the DS group. In the first year of life, the median BVA of the children in the DSN group was 0.1 LogMAR poorer than that of children in the DS group and worsened to approximately 0.2 LogMAR (two lines) poorer beyond the first year of life. Analysis of covariance (ANCOVA) showed a significant difference (F=28.42, p<0.05), with the DSN group having poorer BVA than the DS group.

**Figure 1.**
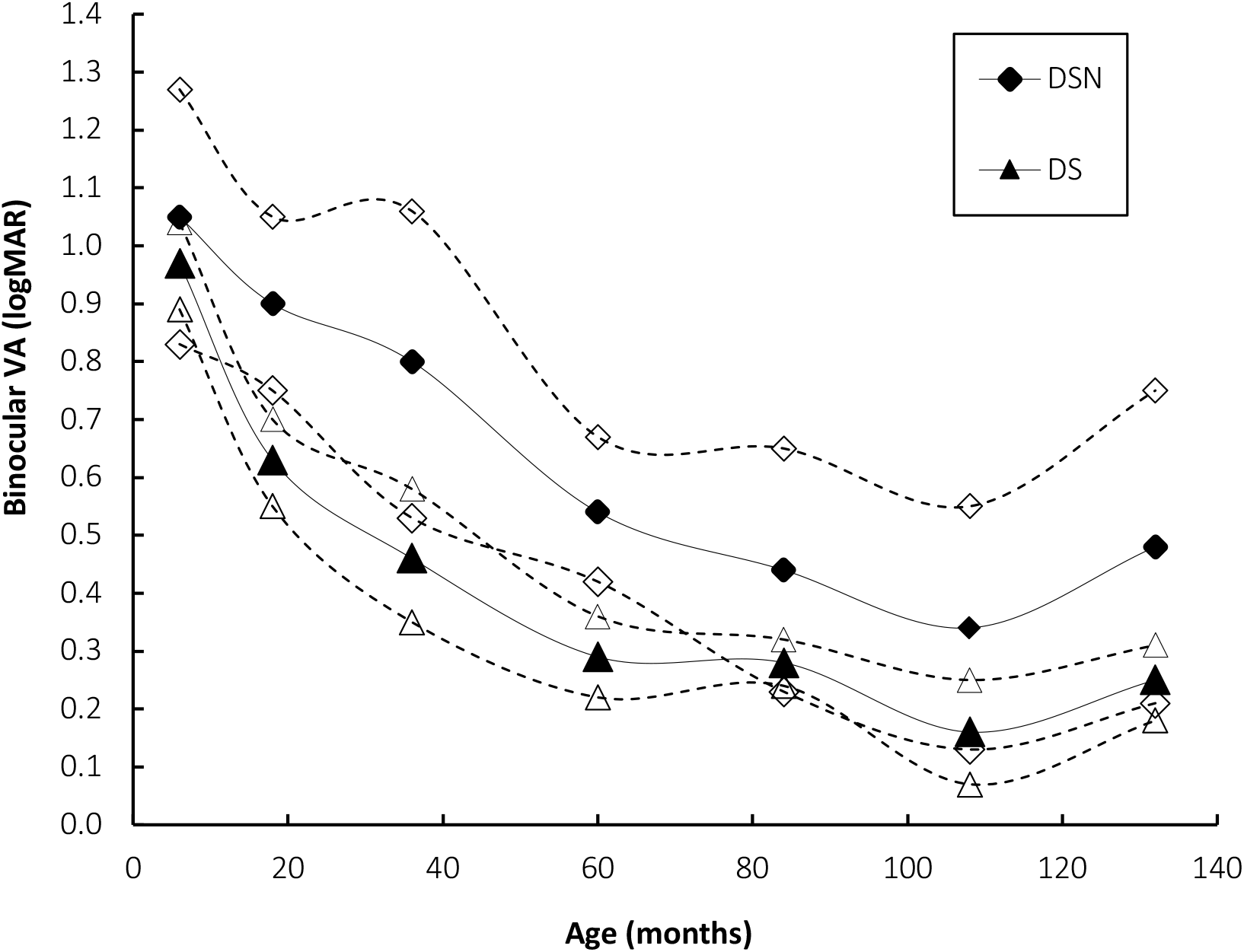
Mean binocular VA of both groups of children with DS, with (DSN) and without (DS) nystagmus. Open markers with dashed lines depict 95% confidence limit

The mean BVA of neurotypical children with idiopathic IN and IN associated with albinism reported by Fu et al.(3) was plotted in Figure 2 along with the mean BVA of children with DS with and without nystagmus from the present study. The BVA of children with DSN is even poorer than that of typically developing children with IN associated with albinism in early childhood but appears better at older ages. The progression of BVA with age of children in the DS group is not markedly different from that of typically developing children with idiopathic IN.

**Figure 2.**
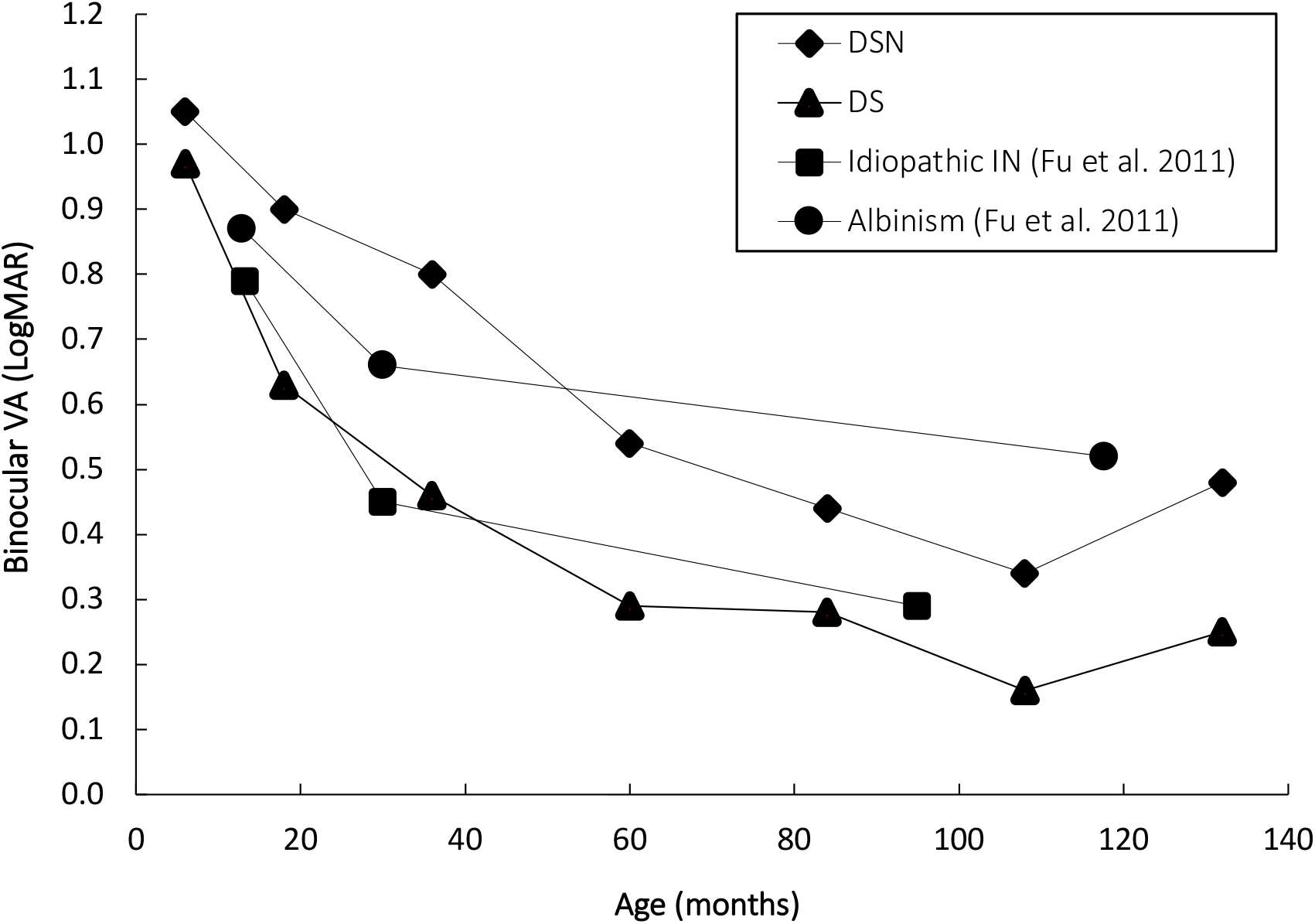
Mean binocular VA of children with DS with (DSN) and without (DS) nystagmus compared to published data of idiopathic IN ad IN associated with albinism (Fu et al. 2011)

### Spherical Equivalent Refractive Error (SER)

Seven (14%) and 18 (10.2%) of children in the DSN and DS group, respectively, were anisometropic. Data of the LE were used for 5 children in the DSN group and 8 children in DS group; otherwise RE data were used. Spherical equivalent refractive error (SER) data of both groups of children were normally distributed (p>0.05) for all age groups except the 12-23.9 months and 10-11.9 years in the DSN group and the 12-23.9 months in the DS group (p<0.05 in all cases). SER in the DSN group was between -12.00D and +7.75D and, in the DS group, between -10.00D and +10.38D.

Figure 3 shows the median SER of both groups of children for each age group. Data for children under 1 year were removed because refractive error is likely to change in early infancy.(25) Although the data were normally distributed, medians and inter-quartile ranges were used to enable comparisons of the results with those reported by Al-Bagdady, Murphy and Woodhouse (2011)(22) whose data were not normally distributed. Children in the DSN group showed more variability in the SER compared to the DS group. Regression analysis showed no significant change in SER with age for both groups of children (DSN, p=0.936; DS, p=0.889). ANCOVA showed a significant difference in the SER between children in the DSN and DS group when age was taken into account (F=8.30, p<0.05).

**Figure 3.**
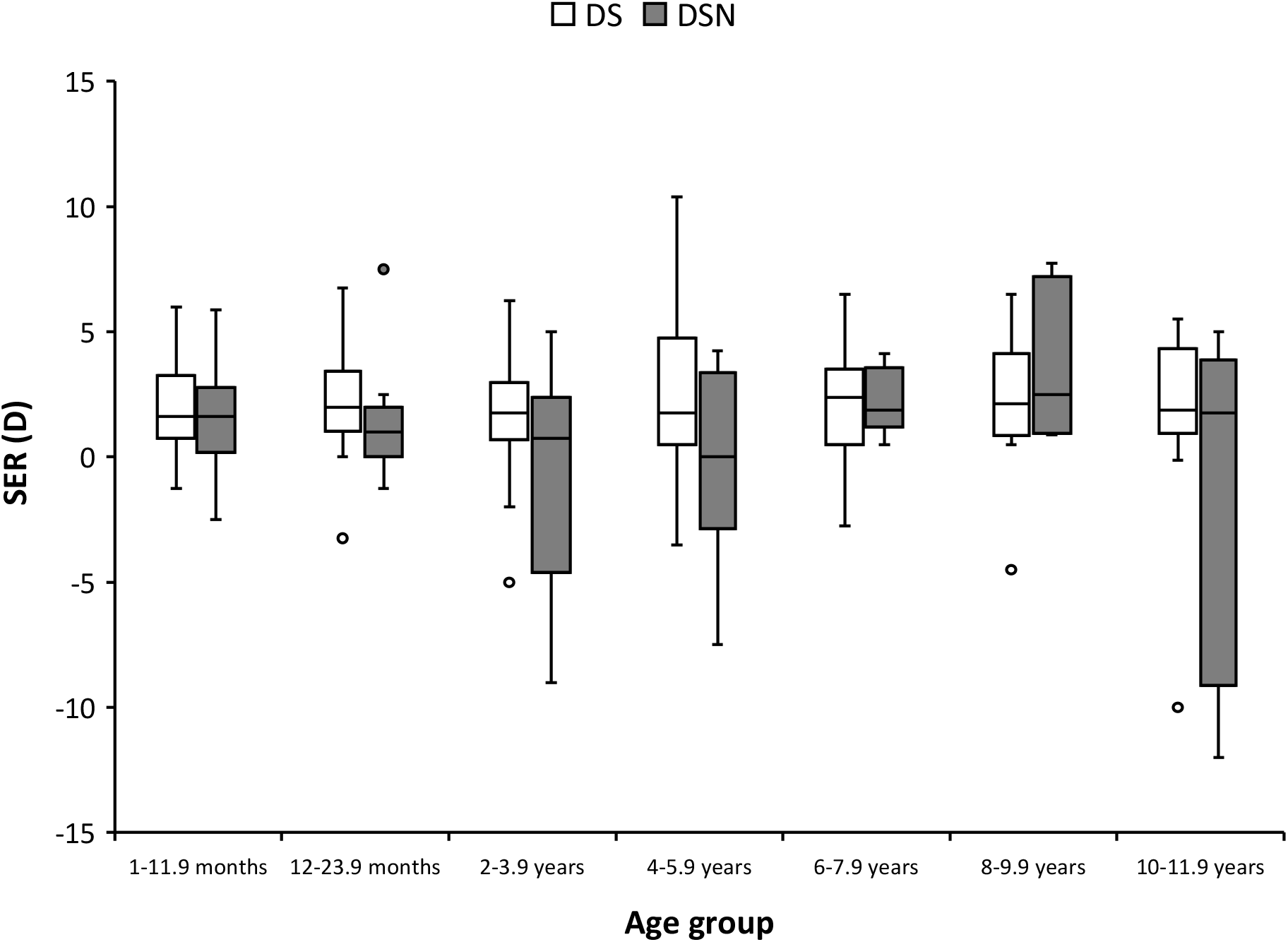
Spherical equivalent refractive error (SER) for each age group of children with DSN and DS. Box represents median and inter-quartile range (IQR). Whiskers represent minimum and maximum values excluding outliers. Circles represent outliers.

The SER distribution of children in the DSN group was plotted alongside SER data of typically developing children with idiopathic infantile nystagmus (IIN) reported by Healey et al.(4). As shown in Figure 4, only 8 (16%) of the children in the DSN group fell outside the 95% confidence limits of the SER of children with DS, and IIN; 7 of these were more myopic and only 1 more hypermetropic.

**Figure 4.**
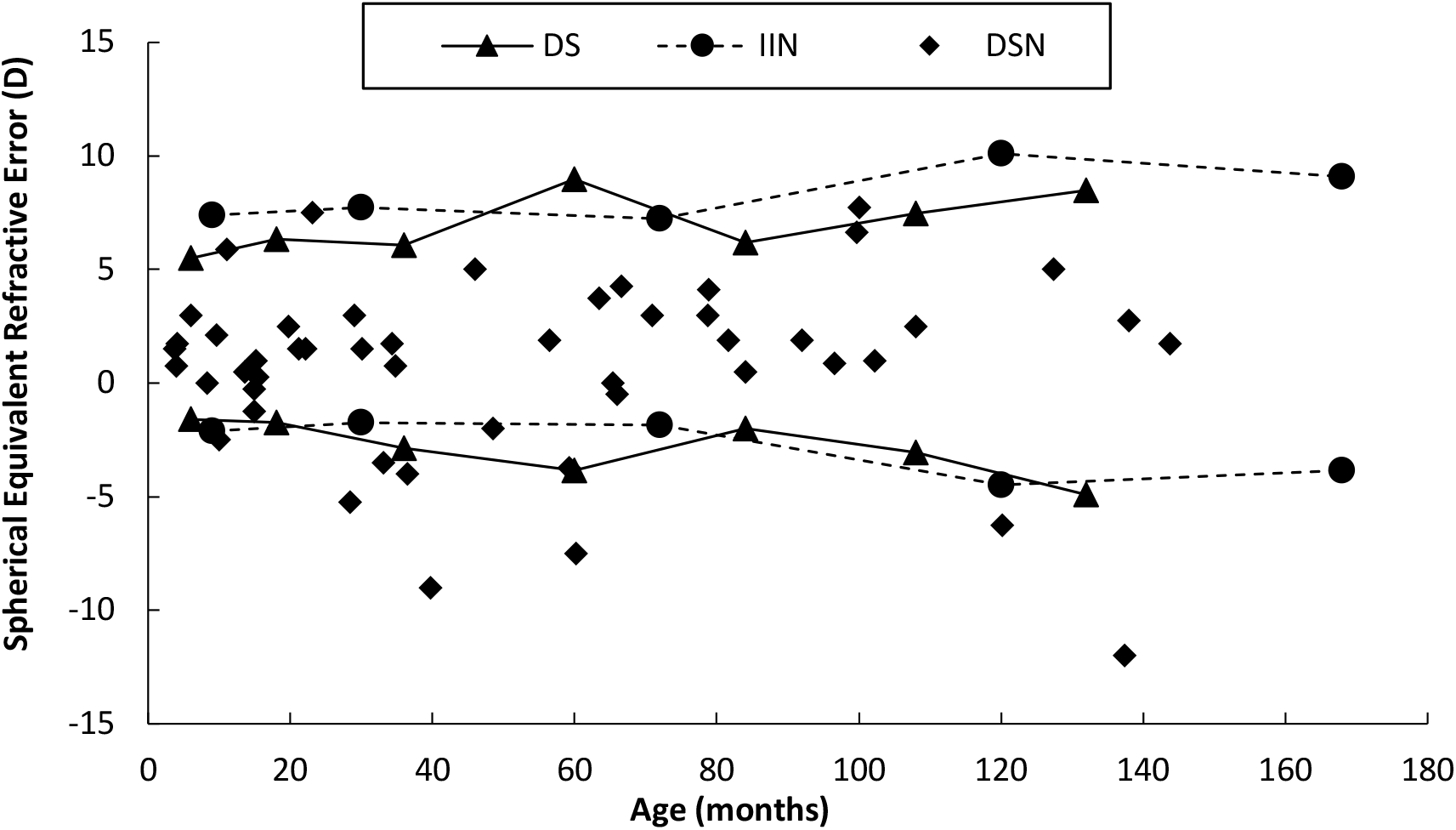
Spherical equivalent refractive error (SER) of children in the DSN group plotted alongside 95% confidence intervals of SER data of children in the DS group (solid lines) from the present study and children with idiopathic infantile nystagmus (IIN, dashed line lines) published by Healy et al. (2014)

Table 2 shows the frequency of type of refractive error for each group of children with DS. Children in the DSN group showed a significantly higher prevalence of myopia (40.7%) than the DS group (11.2%) (χ2= 13.790, p<0.05).

**Table 2.** Frequency of refractive error type in each group of children with DS

|  | Frequency of refractive error type |  |  |
| --- | --- | --- | --- |
|  | Myopia | Hyperopia | Emmetropia |
| DSN (n=50) | 12 (24%) | 14 (32%) | 24 (48%) |
| DS (n=176) | 12(6.82%) | 62(35.23%) | 102 (57.95%) |

### Astigmatism

Mean astigmatism was -0.76 ± 0.62 SD for the children in the DSN group and -0.74 ± 0.81SD for the children in the DS group, which were not significantly different (ANCOVA F= 0.16, p= 0.68). Twenty-seven (54%) of in the children in the DSN group and 102 (58%) children in the DS group had significant astigmatism, a difference that was also not significant (χ2=1.65, p=0.69). Moreover, there was no significant difference in the axis of astigmatism between children with DS and DSN (χ2=1.46, p=0.48).

## DISCUSSION

This retrospective study is the first to exclusively describe the visual and refractive status of children with Down’s syndrome and nystagmus (DSN), who comprise over 20% of our study cohort. The current analysis shows that children with DSN have significantly poorer VA compared to that of children with DS. A recent retrospective analysis of VA data by Zahidi et al.(11) confirms that children with DS have poorer corrected acuity compared to typically developing children by approximately 0.2 LogMAR. The additional deficit in the median acuity of children with DSN was a further 0.2 LogMAR. Felius et al.(5) also reported a VA deficit of 0.4 LogMAR in children with DSN compared to typical norms and suggested that nystagmus is not the sole cause of poor vision in children with DSN. The current study confirms this, as children with DS and no nystagmus appear to have VA on a par with neurotypical children with nystagmus. The difference between typical children and those with DS is not yet fully explained.

The VA deficit of children with IN has been associated with the onset of binocular visual deprivation and the change in the nystagmus waveform.(26) Typically developing children with IN usually present with triangular waveforms during infancy, which transforms into pendular and then to jerk waveforms.(27)(28)(29) This change in the waveform type results in or perhaps is a consequence of changes in the foveation strategy, which underlies the visual performance in IN.(26) Reinecke et al.(27) speculates that children with IN adopt new foveation strategies as they try to focus on the objects that interest them, hence, produces the observed change in the nystagmus waveform type. Therefore, the onset of change from pendular to jerk nystagmus is crucial to the visual development of children with nystagmus. Longitudinal data from infancy were available for only 4 children with DSN, so it is difficult to estimate the timing of the onset of nystagmus for this group of children. Longitudinal eye movement recording data would be needed to determine any changes in the nystagmus waveforms.

One of the limitations of the current study is that VA was measured with current spectacles and not necessarily with the children’s best correction, as testing acuity while wearing a trial frame can distract the children, which may affect their performance during the test. This would apply, of course to both groups. Although there could have been some changes in refractive error since their last clinical correction, this is likely to be small because the children were seen at regular intervals. The visual acuity of the typically developing children with INS in the study by Fu et al.(3) was also measured using ‘habitual optical correction’ as well as with age-appropriate tests. Children with DS have been reported to have poorer VA than the expected norm(6)(8)(9)(10), despite refractive errors being corrected. None of the reported studies measured acuity with the children’s best correction, yet more than 85% of VA in DSN fell below the 95% confidence limits of typically developing children with INS and children in the DS group.

Refractive error of children with DSN differs significantly from that of children with DS who do not have nystagmus over the age of one year old. Although both groups of children were hypermetropic since early infancy, children with DSN were more likely to be myopic compared to the DS group and showed a larger variability in refractive error. Previous studies of children with DS have shown an association between congenital heart defect and nystagmus(30)(31), and between congenital heart defect and myopia(30), and the present study appears to confirm these findings. Neuro-typical children with IN also have been shown to have unconventional refractive development(4), but the similarity in refractive distribution between neuro-typical children with IN and children with DS and no nystagmus is both intriguing and unexplained.

The pattern of the development of astigmatism in children with DS has been reported to differ significantly from typically developing children with no ocular conditions.(21)(22) Children with DS present with WTR astigmatism from an early age, which then develops into oblique astigmatism later during their childhood. On the other hand, neurotypical children with IIN have been reported to present with WTR astigmatism throughout their childhood.(23)(32) Our analysis did not show that nystagmus had any effect on the type of astigmatism in children with DS.

## Data Availability

The data that support the findings of this study are available from the corresponding author JMW upon reasonable request

## SUMMARY

The results of this study have shown that children with DSN have poorer VA than children with DS, in a similar manner to neuro-typical children with nystagmus having poorer acuity than children without nystagmus. However, it is quite clear that there is a significant baseline deficit in acuity attributable to DS, and an additional deficit associated with nystagmus. There was no difference in astigmatism between children with DSN and DS, which is in contrast to typically developing children who have a higher prevalence of astigmatism compared to their counterparts without nystagmus. Finally, the findings of this study show that myopia is associated with nystagmus in children with DS.

It is clear that children with DSN have a visual impairment. It is essential, therefore, that nystagmus in Down’s syndrome receives the same level of attention as it does among typical children, with appropriate advice for parents and targeted educational support for children.

## ACKNOWLEDGEMENTS

The authors would like to thank the past members of the Down’s Syndrome Vision Research Unit, for their contribution to the data collection: Val Pakeman, Mary Cregg, Ruth Stewart, Mohammed Al-Bagdady, Stephanie Campbell and Valldeflors Vinuela-Navarro. Our most important thanks must go to the children and their families for their unfailing support for our study.

